# Quality Improvement Based Implementation and Evaluation of a Decision Aid for Patients with Nephrolithiasis

**DOI:** 10.64898/2026.06.12.26355535

**Authors:** Austin Lee, Saam Kazemi, Patrick Wilson, Karan Thaker, Lorna Kwan, John Cabri, Kevin Li, Matthew Dunn, Alan Yaghoubian, Fuad Elkhoury, Kymora Scotland, Christopher Saigal

## Abstract

**Introduction:** Patients with nephrolithiasis face challenges in making a high-quality, preference sensitive decision. Our prior work established feasibility and patient acceptance of a software-based decision aid (DA). The objectives for this study were to identify implementation strategies for the DA in routine care and determine whether DA implementation enhances decisional quality for patients.

**Methods:** New nephrolithiasis patients were recruited from the institution’s Medical Center from June 2018 to April 2024 to receive a software-based pre-visit DA that measured care preferences and used decision analysis to rank treatments. The RE-AIM framework and Plan-Do-Study-Act (PDSA) cycles were used to improve implementation outcomes. Patients completed survey instruments evaluating decisional conflict, shared decision-making, care satisfaction, and treatment choice following their provider visit. These metrics were compared in the DA cohort (n=81) to those in a “usual care” cohort (n=78) with Wilcoxon rank-sum and Chi-square (or Fisher’s exact) tests.

**Results:** Implementation data revealed sustained reach and progressive improvement in fidelity. The DA cohort reported higher decisional quality relative to controls (p=0.003) and reported greater support/advice to make a choice (p=0.005). The DA cohort more often discussed options with their doctor (87.5% vs 69.2%, p=0.005) and were more likely to be promoters of their provider (p<0.001) and health system (p=0.029). The DA cohort was less likely to have switched their treatment preference post-consultation (32.1% vs 71.8%, p<0.001) suggesting greater consistency in decision-making.

**Conclusions:** Software-based DAs in nephrolithiasis can mitigate decisional conflict, improve SDM, and improve patient satisfaction. Further work should explore broader implementation and long-term clinical outcomes.

## INTRODUCTION

Nephrolithiasis, or kidney stone disease, affects approximately 1 in 11 Americans, with incidence rising over recent decades and recurrence rates reaching as high as 50% within ten years of an initial episode.^1^ Management decisions are often preference sensitive and multifactorial, balancing treatment efficacy, invasiveness, cost, and potential for recurrence or complications.^2^ Available treatment options—including extracorporeal shock wave lithotripsy (ESWL), ureteroscopy with laser lithotripsy (URS/LL), and percutaneous nephrolithotomy (PCNL)—may be equally appropriate depending on stone size, location, and symptoms, and when evaluated against patient preferences for outcomes and potential complications. This preference-sensitive decision occurs in a context of variable patient literacy and health information of uneven quality in public domains.^3^ Such a context is ripe for support with a structured, evidence-based shared decision-making (SDM) process.

SDM has emerged as a valuable approach in many preference-sensitive decision contexts, empowering patients to make informed choices aligned with their values.^4–6^ Multiple studies have demonstrated that SDM can improve patient satisfaction, reduce decisional conflict, and even decrease unnecessary interventions.^7–9^ Despite these benefits—and growing policy-level endorsements of SDM including within the Affordable Care Act— its implementation in routine urologic care remains inconsistent.^10–12^

One promising strategy to standardize and facilitate SDM is the deployment of decision aids (DAs), which present tailored, evidence-based information about options and outcomes to patients. DAs have been shown to enhance knowledge, clarify personal values, and reduce decisional conflict across a variety of surgical and chronic care contexts.^5,13^ Electronic decision support systems, in particular, offer scalable, asynchronous delivery and can integrate preference elicitation and personalized output for both patients and providers.^14,15^ Yet even among validated DAs, real-world uptake remains limited due to practical barriers in clinical workflows, variable patient engagement, and concerns about sustainability.^14^

To address both the implementation challenges and clinical impact of SDM tools in urology, we deployed a software-based decision aid (WiserCare, Inc. 2017) within routine nephrolithiasis care at a large academic health system. The DA was designed to be delivered asynchronously before consultation and used a personalized decision analysis model built “on the fly” for patients. The model uses preference measurement through conjoint analysis to provide weights and generates a rank list ordered on evidence-based treatment options for the patient to review prior to the counseling visit. We hypothesized that using the framework and tools of implementation science, we could deploy and scale this intervention successfully in our health system.

Our study objectives were twofold: first, to identify strategies to improve the implementation of this DA in routine care, and second, to determine whether DA implementation enhances decision quality for patients facing newly diagnosed kidney stones.

## METHODS

### Study Design Overview

We conducted a multi-stage implementation science study at UCLA Health from June 2018 to April 2024 to assess the implementation and impact of a decision aid (DA) for patients newly diagnosed with nephrolithiasis (**Figure 1**). It was paused for a period around the COVID pandemic. The project consisted of two overlapping but distinct components: (1) A quality improvement (QI) initiative guided by the RE-AIM framework^16^ and using Plan-Do-Study-Act (PDSA) cycles focused on improving implementation outcomes. (2) An observational cohort study comparing patient-reported decisional outcomes between those who received the DA versus those who received usual care (pre-post design).

**Figure 1.**
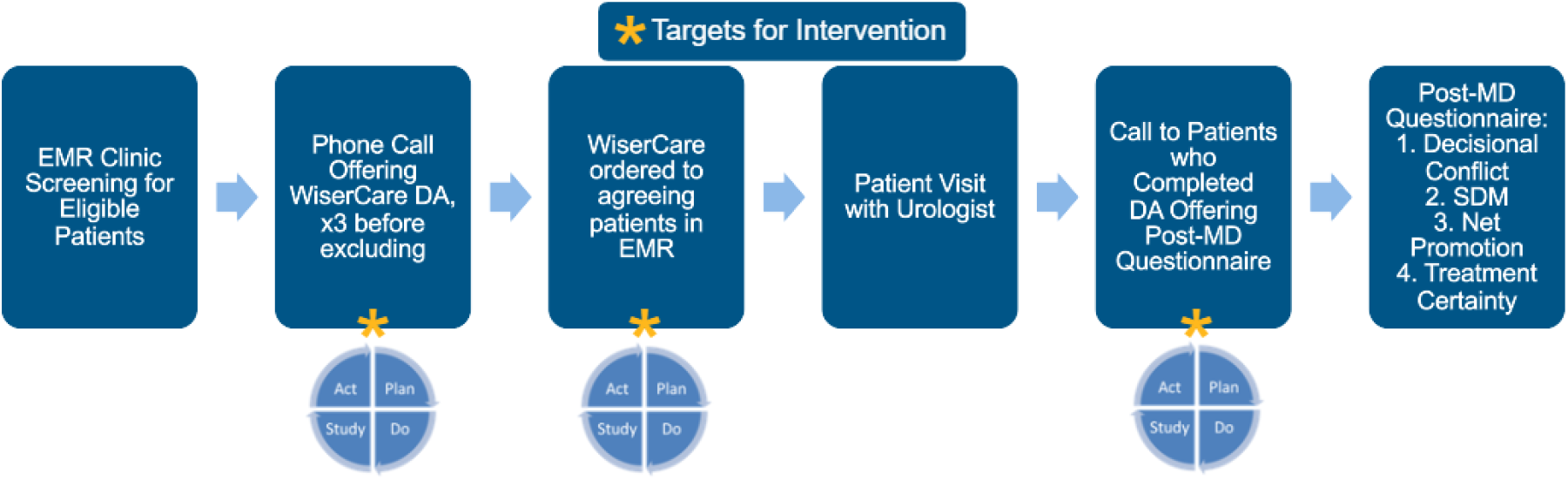
Patient Flow Diagram. Patient flow diagram illustrating implementation of the WiserCare decision aid (DA). Eligible patients were identified via EMR, contacted by phone, and assigned the DA if they consented. Post-consultation, patients who completed the DA were surveyed on decisional conflict (SURE), shared decision-making (SDM), Net Promoter Score (NPS), and treatment certainty.

### Setting and Participants

The study was conducted across general and endourology practices within UCLA Urology. Patients were eligible if they were: age ≥18, English-speaking, presenting for a new evaluation of kidney or ureteral stones, and had imaging-confirmed nephrolithiasis (based on CT, ultrasound, or external documentation).

Patients with previously treated stones were eligible if new, untreated calculi were documented. Exclusion criteria included lack of access to UCLA’s patient portal, prior urologic consultation for the same stone episode, or history of dementia or other neuropsychiatric disorders that would preclude participation.

### Intervention Group: WiserCare Decision Aid

Eligible patients were sent a secure link to the WiserCare Kidney Stones module via the MyChart patient portal 1–7 days prior to their urology consultation. The DA included:

1. Educational material about nephrolithiasis and treatment options
2. Preference elicitation using a conjoint analysis interface
3. A personalized “best-fit” treatment recommendation based on a personalized decision analysis incorporating a patient’s predicted outcomes and stated preferences

The output report was shared with both the patient and provider via the EMR before the visit.

### Control Group: Usual Care

The control group consisted of a historical cohort of 78 patients surveyed from March 2016 to May 2017 who underwent standard urology consultation for nephrolithiasis prior to implementation of the WiserCare DA. These patients did not receive the DA but completed the same post-consultation survey as the intervention group.

### Implementation Outcomes

We used the RE-AIM framework to evaluate the following implementation outcomes:

**Reach**: Proportion of eligible for the DA who were successfully identified and invited to complete the DA

**Fidelity**: Proportion of invited patients who completed the DA as intended

Implementation performance was evaluated using Statistical Process Control (SPC) charts^17^ to track decision aid (DA) reach and fidelity over time. Reach was evaluated by comparing new patient visit lists, and WiserCare invitation logs. The new patient lists were recorded internally within REDCap after reviewing patient charts to confirm new stone visits against scheduler notes. Fidelity was measured by evaluating WiserCare data on the number of patients who completed a module versus the number who were invited to complete the module. We conducted iterative Plan-Do-Study-Act (PDSA) cycles to improve our implementation outcomes.

### Effectiveness Outcomes

To evaluate effectiveness of the DA, we compared patients exposed to the DA (intervention group) with the historical cohort who received usual care (control group). We measured decision quality, conceptualized as a basket of related measures.

#### Primary Outcome

- Presence or absence of decisional conflict, measured using a dichotomized version of the validated 4-item SURE scale (absence (SURE score 4) vs presence (SURE score 0-3))^18^

#### Secondary Outcomes

- Decisional Conflict Scale^19^
- Perceived shared decision-making, measured by select items from the SDM-Q^20^
- Net Promoter Score (NPS) for physician and health system^21^
- Satisfaction with decision and willingness to adhere to treatment

### Data Collection

Patients completed post-consultation survey decision quality instruments online or in person within 1 week after their consultation, including designation of their post-visit treatment preference. We also collected ultimate treatment choice, which was abstracted from the medical record.

### Statistical Analysis

For implementation outcomes, SPC charts were used to visualize trends and shifts in performance relative to control limits, after PDSA improvement cycles. Standard SPC rules were applied to differentiate between common cause and special cause variation. Special cause variation was identified using established rules, including shifts of ≥8 consecutive points on one side of the center line, trends of ≥6 increasing or decreasing points, or points outside control limits. When special cause variation was detected, center lines and control limits were recalculated to reflect the new process baseline. PDSA cycles were iteratively conducted every week to optimize DA reach and fidelity over time.

Continuous variables were compared using t-tests (or Wilcoxon rank-sum tests) and categorical variables (e.g., dichotomized SURE score, treatment switching, SDM perception) were assessed with the Chi-square (or Fisher’s exact) test, as appropriate. A p-value <0.05 was considered statistically significant.

### Ethics

This study was approved by the UCLA Institutional Review Board (#19-000872). The implementation component was conducted under a QI designation; the impact analysis was approved as non-exempt human subjects research.^3^

## RESULTS

From June 2018 to April 2024, 81 patients completed both the WiserCare DA and the post-consultation survey and were included in the intervention cohort.

### Cohort Characteristics

For the clinical impact assessment, 159 patients were included: 81 in the intervention group (received the DA) and 78 in the control group. The intervention group was significantly older (mean age: 57.0 vs. 51.6 years, *p*=0.033) (**Table 1**). Racial/ethnic distribution differed between groups (*p*=0.001), with a higher proportion of Asian and a lower proportion of non-Hispanic white patients in the intervention cohort. All other demographic (e.g. sex, relationship status, and employment status) and clinical characteristics were similar across groups (all *p*>0.05).

**Table 1.** Patient Characteristics (N=159). Demographic and clinical variables for the control (n=78) and intervention (n=81) groups.

|  | Total<br>(N=159) | Control<br>(N=78) | Intervention<br>(N=81) | P-value |
| --- | --- | --- | --- | --- |
| **Age at Qx, Mean (SD)** | 54.2 (15.63) | 51.6 (15.77) | 57.0 (15.10) | 0.0330<sup>1</sup> |
| **Sex, n (%)** |  |  |  | 0.3544<sup>2</sup> |
| Female | 60 (39.5%) | 28 (35.9%) | 32 (43.2%) |  |
| Male | 92 (60.5%) | 50 (64.1%) | 42 (56.8%) |  |
| **Race/Ethnicity, n (%)** |  |  |  | 0.0014<sup>3</sup> |
| Asian | 17 (11.3%) | 5 (6.4%) | 12 (16.7%) |  |
| Black or African American | 3 (2.0%) | 1 (1.3%) | 2 (2.8%) |  |
| Hispanic | 13 (8.7%) | 5 (6.4%) | 8 (11.1%) |  |
| Non-Hispanic | 88 (58.7%) | 53 (67.9%) | 35 (48.6%) |  |
| Other | 21 (14.0%) | 14 (17.9%) | 7 (9.7%) |  |
| Refused | 8 (5.3%) | 0 (0.0%) | 8 (11.1%) |  |
| **Relationship Status, n (%)** |  |  |  | 0.1638<sup>3</sup> |
| Married | 103 (68.2%) | 49 (62.8%) | 54 (74.0%) |  |
| Not Married | 48 (31.8%) | 29 (37.2%) | 19 (26.0%) |  |
| **Employment Status, n (%)** |  |  |  | 0.8627<sup>3</sup> |
| Employed | 97 (65.5%) | 52 (66.7%) | 45 (64.3%) |  |
| Not Employed | 51 (34.5%) | 26 (33.3%) | 25 (35.7%) |  |

<sup>1</sup>Equal variance two sample t-test; <sup>2</sup>Chi-Square p-value; <sup>3</sup>Fisher Exact p-value;

### Implementation Outcomes

Weekly SPC data—our primary *learning measure*—revealed sustained reach (the proportion of eligible patients who received the DA) and progressive improvement in fidelity (the proportion who completed it as intended), with one point of special cause variation for fidelity triggering center line adjustment (**Figure 2**). Fidelity improved from 33% to 48%. Reach remained in control.

**Figure 2.**
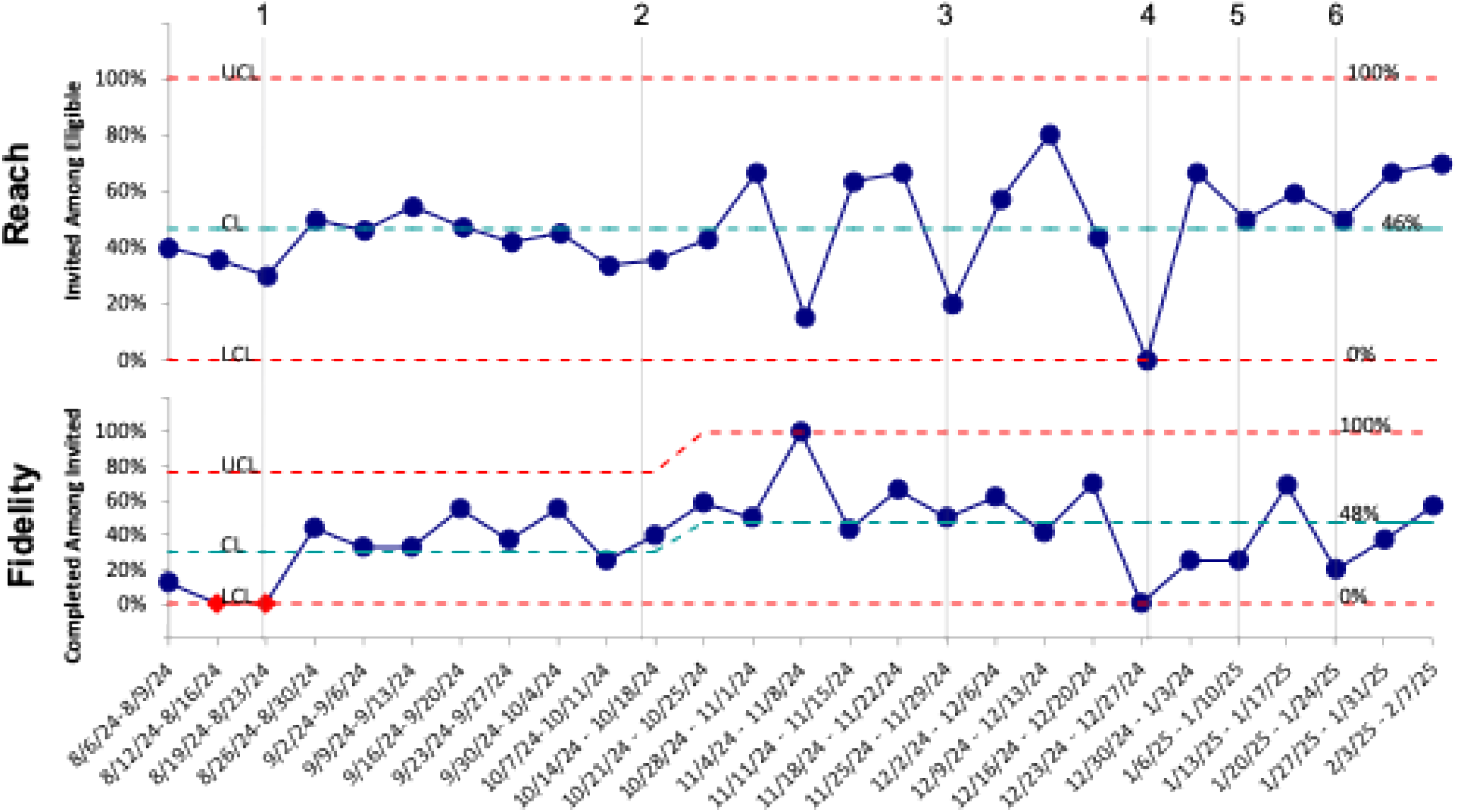
Statistical process control (SPC) charts illustrating decision aid (DA) reach, and DA fidelity over time. Key events are annotated: (1) Broken link to DA module identified and fixed (2) Survey of patients who were reached but did not complete the module led to standardized call script (3) Thanksgiving holiday (4) Winter holiday (5) Los Angeles wildfires (6) Launch of in-person recruitment

Key quality improvement interventions included fixing a broken DA hyperlink, standardizing outreach scripts, and initiating in-person recruitment, all of which coincided with upward shifts in performance in fidelity. Periodic declines in metrics were aligned with known external disruptions (e.g., holidays, wildfires).

To better understand the emerging *capacity* of the implementation system over time, we also plotted monthly performance trends (**Figure 3**). While longer-term data will be required to determine the stability and upper performance limits of the system, initial monthly trends suggest improved reliability in DA delivery and completion.

**Figure 3.**
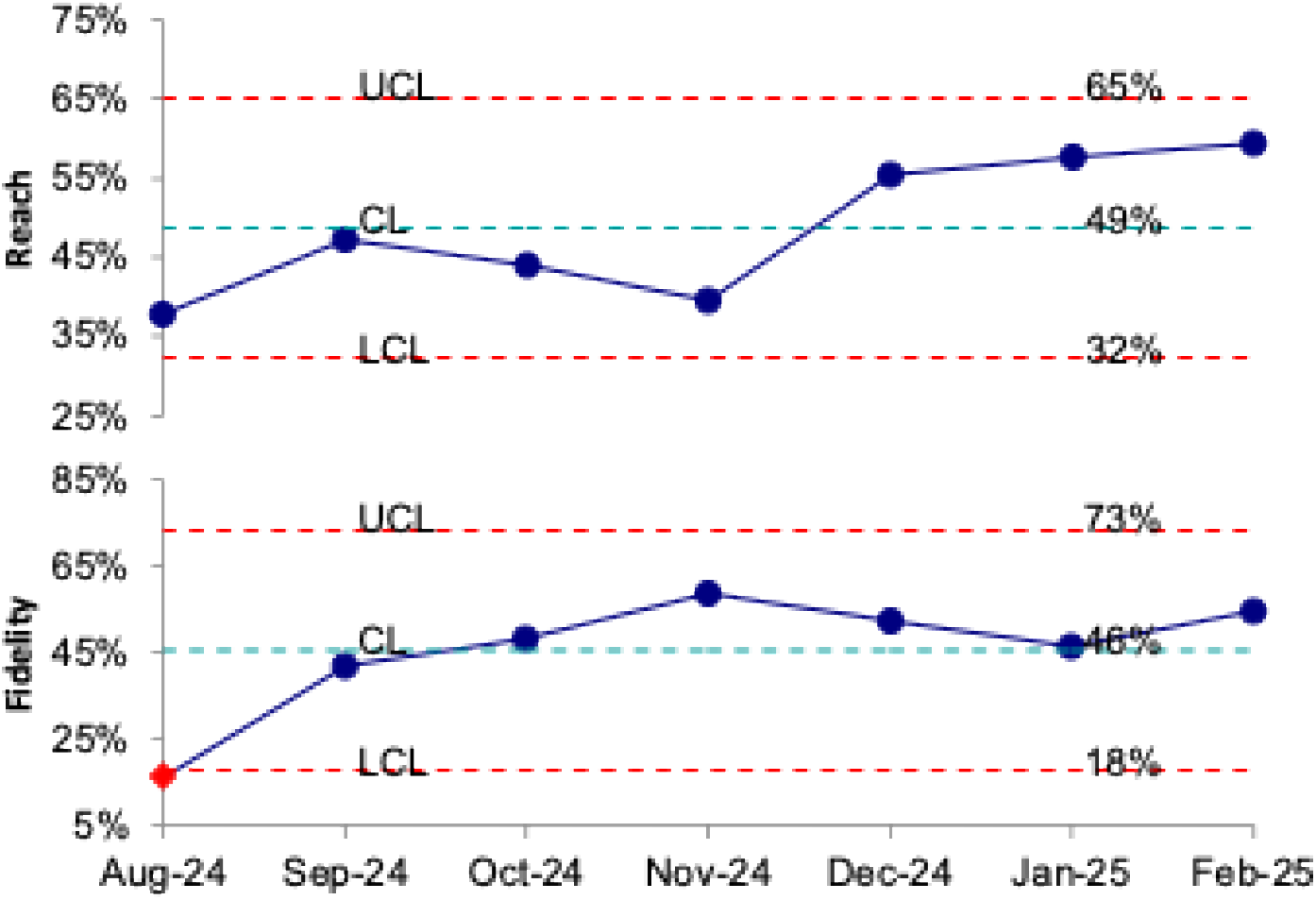
Monthly statistical process control (SPC) charts depicting (A) decision aid (DA) reach and (B) DA fidelity over time. Dashed red lines represent upper and lower control limits. Dashed cyan line indicates baseline center line. Data points reflect monthly aggregation of performance metrics, offering a high-level view of system capacity beyond short-term fluctuations.

### Decisional Outcomes

Patients in the intervention group reported higher decision quality relative to controls (**Table 2**). While absence of decisional conflict was not significantly different (65.4% vs. 53.8%, *p*=0.136), DA recipients scored higher in a specific area of decision quality. They were more likely to report knowing the benefits/risks of treatment (84.0% vs. 62.8%, *p*=0.003). They also reported better shared decision-making where they more often discussed options with their doctor (87.5% vs. 69.2%, *p*=0.005) and were more likely to be Net Promoter Score “promoters” for their physician (90.9% vs. 55.8%, *p*<0.001) and for the health system (85.9% vs. 67.9%, *p*=0.029).

**Table 2.**
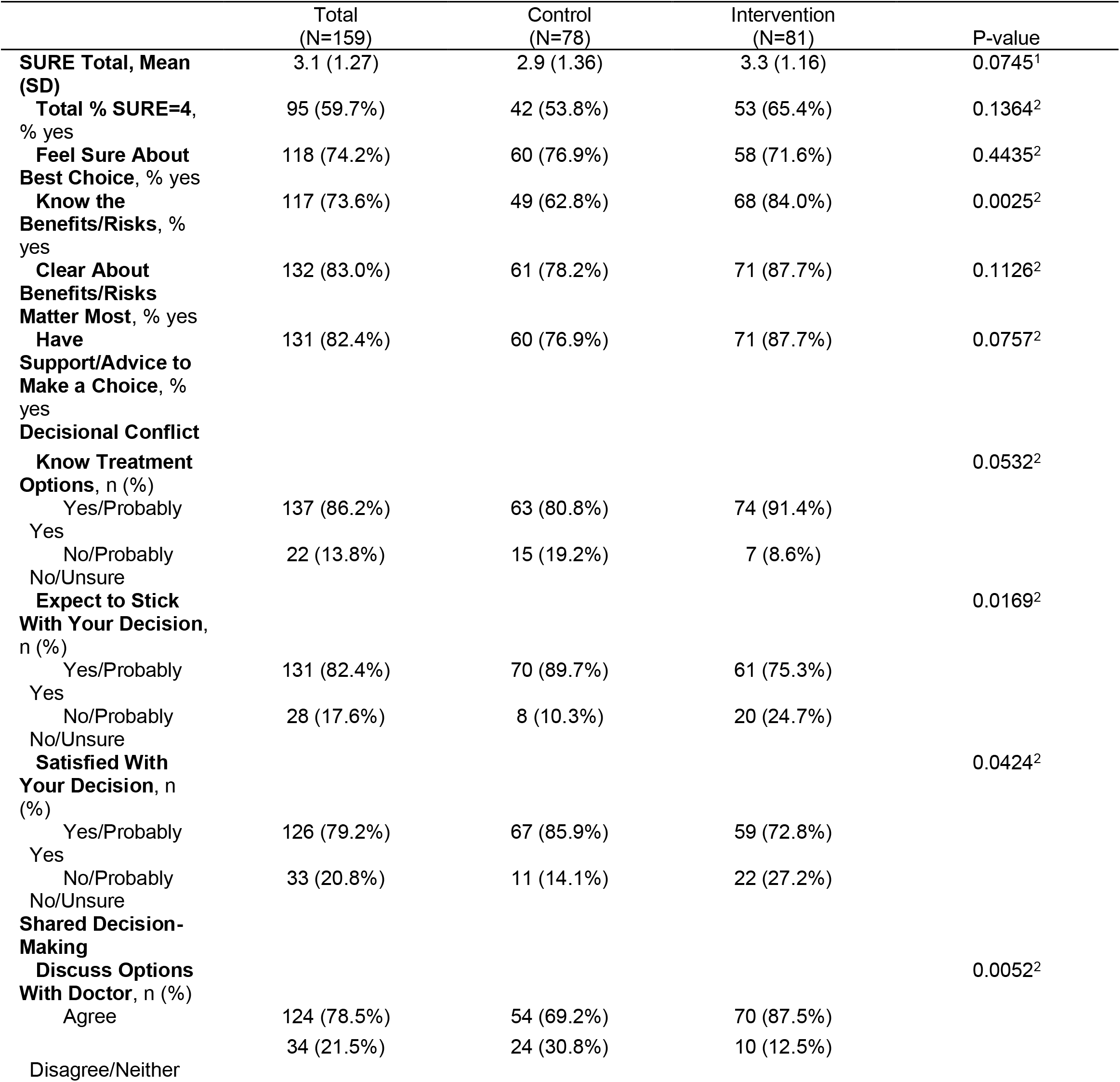
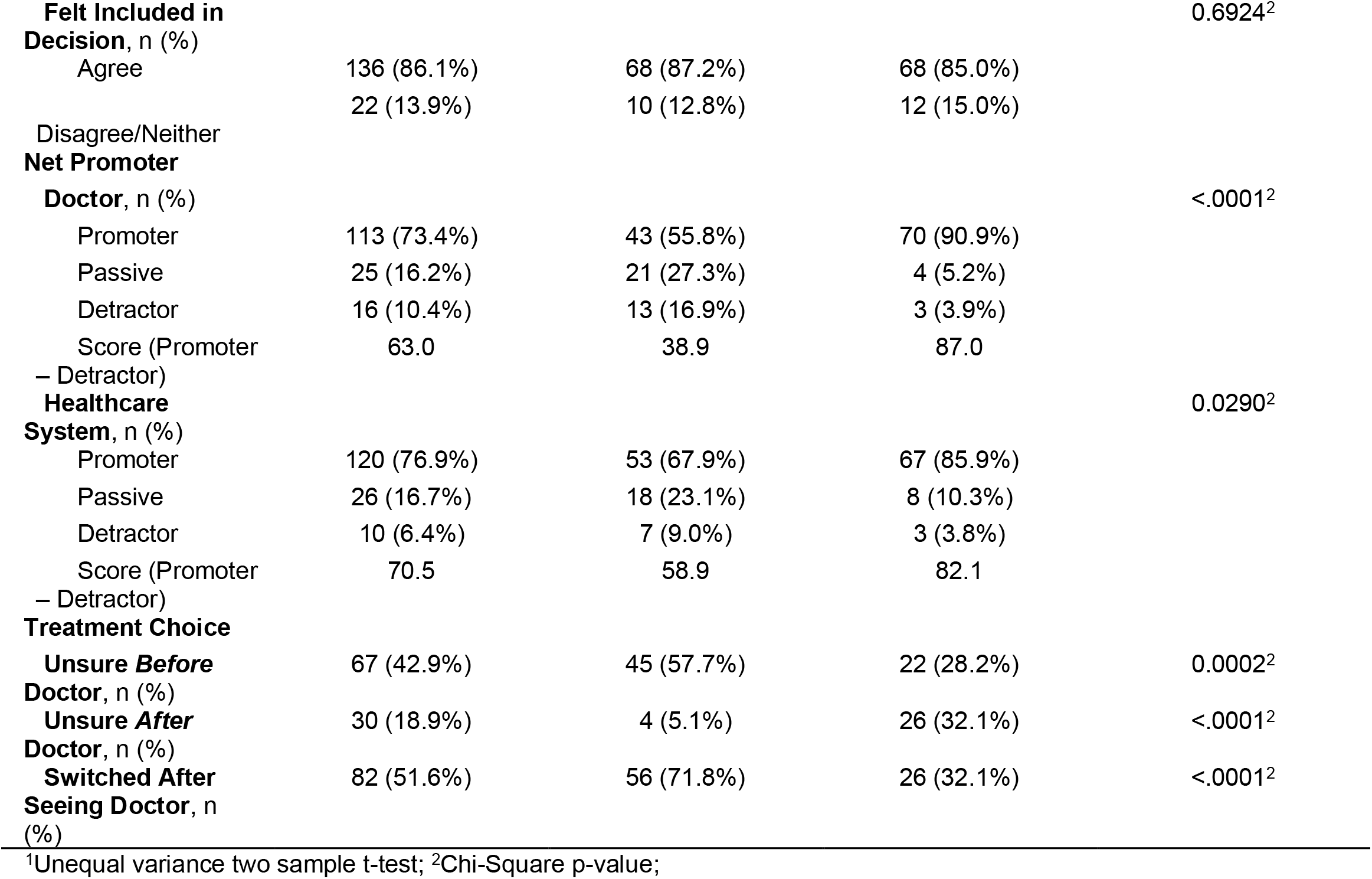
Summary statistics for SURE, decisional conflict, shared decision-making, net promoter, and treatment decisions (N=159) Summary of Decision Quality and Related Outcomes. Comparison of control (n=78) and intervention (n=81) groups on SURE scores, decisional conflict indicators, shared decision-making perceptions, Net Promoter Scores, and treatment-related behaviors. Results presented as means (SD) or percentages (% yes). P-values reflect t-tests or chi-square tests as appropriate.

Although DA patients were more likely to be unsure of their treatment preference after seeing the physician (32.1% vs. 5.1%, *p*<0.001), they were less likely to have switched choices post-consultation (32.1% vs. 71.8%, *p*<0.001), suggesting greater consistency in their decision-making process. Of note, uncertainty may have been driven by clinical factors as 14 out of 26 (53.8%) DA patients who were unsure of their treatment preference post-MD visit had actually either passed their stone prior to the appointment, or were advised to undergo additional imaging prior to making a treatment decision at a future appointment. A further 2 out of 26 patients had special external factors impacting their decision-making which may not be captured by the DA (one was recommended “prophylactic” URS to avoid complications during upcoming travel, the other presented with a wound vac and plan for eventual skin graft prior to stone intervention).

DA recipients who remained unsure of their treatment preference post-MD visit reported higher NPS (doctor and health system) than those who were sure then became unsure. Interestingly, 5 out of 12 (41.7%) patients who switched from sure to unsure of their treatment preference after seeing the physician still reported a SURE score of 4.

DA patients were also less likely to report satisfaction with their decision (72.8% vs. 85.9%, *p*=0.042) and expectation to adhere to their choice (75.3% vs. 89.7%, *p*=0.017).

## DISCUSSION

In this multi-stage study, we implemented and evaluated a shared decision-making (SDM) decision aid (DA) for nephrolithiasis care within a large academic urology practice. Our findings demonstrate that electronic DAs can be feasibly integrated into clinical workflows and may improve multiple dimensions of decision quality.

Although overall SURE scores showed no significant improvement, specific elements of decisional confidence—particularly understanding of risks and benefits—were significantly higher among patients exposed to the DA. This aligns with prior literature emphasizing the value of structured educational tools in enhancing patient knowledge and clarity of values.^1,5^

Engagement in shared decision-making was meaningfully elevated in the intervention group, as evidenced by greater discussion of options with providers. Importantly, patients who used the DA were substantially more likely to endorse their physician and healthcare system via Net Promoter Scores, reinforcing the broader relational and reputational benefits of well-executed SDM strategies.^21^

Interestingly, DA recipients were more likely to remain unsure about their treatment preference after seeing their physician, yet they were also significantly less likely to change their treatment choice post-consult. This paradox may reflect a more deliberate and informed decision-making process, where patients maintain flexibility while ultimately adhering to a consistent plan aligned with their values. It is also noted that the majority of DA recipients who felt uncertainty after their consultation were potentially impacted by dynamic clinical factors, such as stone passage or recommendation for specialized imaging, which would not necessarily be captured at the time of DA completion.

Notably, the intervention group showed lower satisfaction and adherence expectations— an unexpected finding that may suggest increased decisional awareness prompting more critical reflection, a phenomenon previously described in SDM research.^4,7^ Indeed, DA recipients who remained unsure in their treatment preference were still more likely to be Net Promoter Score “promoters” for their physician and healthcare system than DA recipients who changed their preference after consultation, suggesting they felt informed and supported in their decision-making. Furthermore, multiple DA patients who switched from sure to unsure after their consultation still scored highly on the SURE scale, which again may suggest increased reflection on treatment choices rather than true decisional conflict. It is possible that the DA was unable to capture other clinical and social factors (upcoming travel etc.) that may have impacted some of the patient decisions.

Our implementation strategy—leveraging RE-AIM and PDSA cycles—successfully improved DA fidelity over time. However, persistent variation in reach suggests that broader scale-up will require additional focus on workflow integration, patient engagement strategies, and clinician adoption.

The initial cohort of this intervention included English-speaking patients at a single academic center, raising questions of potential access disparities for minority patient populations across resource-constrained public and safety net hospitals. Minority populations experience lower rates of health literacy, limited healthcare resources, and communication barriers with nonminority physicians which may lead to differences in SDM between minority and nonminority patients.^22-26^ However, previous work by our team directly compared WiserCare DA implementation for prostate cancer between academic and public (county) care centers and found that despite significant differences in patient demographic and clinical characteristics, there was no significant difference in implementation (reach and fidelity), decisional quality, or treatment satisfaction between sites.^27^ Of note, targeted implementation strategies such as the creation of a Spanish-translated DA increased completion rate in the county where the eligible minority patients were predominantly Hispanic or Latino.^27^ Taken together, these findings affirm that SDM may broadly benefit patients regardless of demographics or system characteristics and reiterate the importance of tailored efforts in future DA expansion to facilitate equitable access for disadvantaged populations across different systems.

Other limitations include the non-randomized and unblinded design, inherent reliance on unmatched historical controls which may mask the outcome variation between specific consultants, and the modest sample size which may limit generalizability. It is also noted that the particulars of counseling between the historical control and intervention group may vary due to the rapid uptake of new clinic tools including thulium, suction sheaths, and increased usage of mini PCNL. Additionally, selection bias may have influenced who completed the DA, and survey-based outcomes may be susceptible to response bias.

## CONCLUSION

Our findings support the real-world feasibility and value of SDM decision aids in urologic practice. Electronic tools like WiserCare can improve decision confidence, patient-provider communication, and system reputation—offering a scalable path toward more patient-centered kidney stone care. Future studies should explore broader implementation, randomized assignment within a fixed group of providers, and long-term clinical and cost outcomes.

## Data Availability

All data produced in the present study are available upon reasonable request to the authors

